# Adapting Clinical Event Annotation to Dutch Primary Care: An Event Annotation Framework for Post-Acute Infection Syndromes

**DOI:** 10.64898/2026.08.19.26360841

**Authors:** Sara Mazzucato, Artuur Leeuwenberg, Sander van Doorn, Joost van Rosmalen, Isabel Slurink

## Abstract

Clinical natural language processing (NLP) relies on language-specific corpora, which remain scarce for Dutch primary care. This limits the use of NLP to study post-acute infection syndromes (PAIS), such as post-COVID condition, as relevant symptoms and their temporal context are often documented in free-text general practitioner (GP) records rather than structured codes. The COVID-19 Annotated Clinical Text (CACT) framework was developed to identify COVID-19 diagnoses and related symptoms in English-language clinical notes, each represented as an *event*: a text span linked to its context, such as negation, timing and certainty. We translated and adapted CACT for Dutch primary care, extending its scope beyond COVID-19 to PAIS and other relevant infections and diagnoses, with annotation rules tailored to the four sections of Dutch GP notes (SOEP: Subjective, Objective, Evaluation, Plan). The revised framework distinguishes symptoms, acute infections, PAIS and other relevant diagnoses, and captures 11 types of supporting diagnostic evidence, including laboratory tests or physical examinations. It also differentiates uncertain diagnoses (‘possibly post-COVID’) from conditions mentioned but not asserted (e.g. a patient’s fear of long COVID). In a pilot of 200 GP notes independently annotated by two annotators, over-all span-level agreement over six core categories (symptoms, diagnoses, negation, evidence, timing and vital signs) was *F*_1_ = 0.51 (95% CI [0.47, 0.55]). Agreement increased to *F*_1_ = 0.78 [0.75, 0.80] when restricted to text spans marked by both annotators, suggesting that disagreement primarily concerned whether a mention was annotated at all, not which category it received. Future research should evaluate which parts of the revised framework generalise beyond Dutch primary care and PAIS.

## 1 Introduction

Clinical natural language processing (NLP) requires language-specific resources, but most existing tools and annotated corpora have been developed for English-language clinical text (Klug et al., 2024). Dutch biomedical NLP lags behind (Leeuwenberg and Kuiper, 2026). Dutch clinical encoders exist (Bosma et al., 2025; Verkijk and Vossen, 2025) and isolated annotation efforts for hospital records have been published (Homburg et al., 2023), but to our knowledge, no publicly available event-extraction corpus exists for Dutch primary-care text. This gap is particularly relevant for research on post-acute infection syndromes (PAIS), such as post-COVID condition, which are characterised by heterogeneous symptoms, variable trajectories, and diagnostic uncertainty. General practitioner (GP) electronic health records provide valuable longitudinal information on infections, symptoms, diagnoses, and their progression over time, but much of this information is documented in free-text consultation notes rather than structured International Classification of Primary Care (ICPC) codes. Systematic extraction therefore requires NLP methods that can identify clinical concepts together with their temporal and diagnostic context. Dutch GP free-text consultation notes follow the SOEP format, the Dutch equivalent of SOAP, with sections for the Subjective complaint (*Subjectief*), Objective findings (*Objectief*), Evaluation (*Evaluatie*), and Plan (*Plan*). A diagnosis or symptom mention is often difficult to interpret without surrounding context: *post-COVID* is only actionable once tied to onset (‘3 weeks ago’, *sinds 3 weken*), certainty (‘possibly’, *mogelijk*), evidence (‘PCR test positive’, *PCR test positief*), and negation status (‘ruled out’, *uitgesloten*). An event-based annotation approach, in which clinical concepts are represented as triggers linked to contextual arguments, preserves this compositionality and supports down-stream phenotyping. This design, established in clinical concept-and-assertion extraction (Uzuner et al., 2011; Gao et al., 2022), underlies the COVID-19 Annotated Clinical Text (CACT) framework (Lybarger et al., 2021): an event schema representing COVID-19 diagnoses and symptoms as triggers linked to contextual arguments (e.g., Negation, Severity, Assertion), rather than using flat span labels. This allows contextual information to be represented separately from the clinical concept. CACT forms the basis for our adaptation. This work was developed within a broader study of post-COVID and other PAIS in Dutch primary care, for which GP records are a principal data source. Machine translation followed by an English extractor and direct cross-lingual transfer behave very differently on clinical named entity recognition (NER) (Gaschi et al., 2023). However, neither has been validated on Dutch primary-care text, which is typically short, abbreviation-heavy, and SOEP-structured. We translated CACT (Lybarger et al., 2021) and adapted it to Dutch general practice, broadening its scope to clinical event extraction across symptoms, acute infections, diagnoses, and their contextual attributes, while adding specific ex-tensions for PAIS research. Our contributions are:

1. a Dutch clinical event-annotation framework for primary care, covering symptoms, infections, diagnoses, and contextual information, with ex-tensions for post-COVID and PAIS phenotyping;
2. a 200-note pilot annotated independently by two medical students, with inter-annotator agreement (IAA) under three complementary criteria;
3. openly released resources, including the annotation guideline, the INCEpTION schema export and 500 synthetic notes, under CC BY 4.0; and
4. conditional access to the annotated GP corpus for research, which cannot be released openly under GDPR (Ethical Considerations).

## 2 The Annotation Framework

### 2.1 Schema Overview

We used an event-based model (full schema in Table 1, Appendix A) to annotate all symptoms, infections and diagnoses mentioned in the notes, regard-less of their potential relation to PAIS. Each *event* consists of a *trigger span* (the shortest text identifying the clinical phenomenon) and *arguments* capturing its context. Arguments are linked to triggers through relation arcs, allowing contextual information to be annotated separately from the clinical concept itself. For example, in Figure 1, *post-COVID* is annotated as the trigger of a diagnosis event, while *Mogelijk* (‘possibly’) is linked as an argument indicating diagnostic uncertainty. We distinguished two trigger types: Symptom, including patient-reported complaints, examination findings and clinician-observed signs, and Diagnostic-Expression, including diagnoses and conditions, divided into three subtypes. Events can be linked to contextual arguments describing Timing, Negation, Subject, Actuality, Hedged and Severity. Diag-nosticExpression can additionally be linked to an EVIDENCE argument with 11 possible values. Arguments were annotated only when explicitly mentioned or when their value deviates from a documented default (Negation: *present*; Subject: *patient*; Actuality: *actual*; Hedged: *not_hedged*). Thus, in the absence of an explicit modifier, a symptom mention was interpreted as referring to a present symptom in the patient. We additionally annotated Measurement as a separate, non-event construct, consisting of paired *type* and *value* spans (e.g., temperature, SpO_2_, blood pressure) documented in the Objective section. These measurements provide clinical information that may support or contextualise nearby symptoms and diagnoses, while remaining separate from the event-based annotation structure.

**Table 1:**
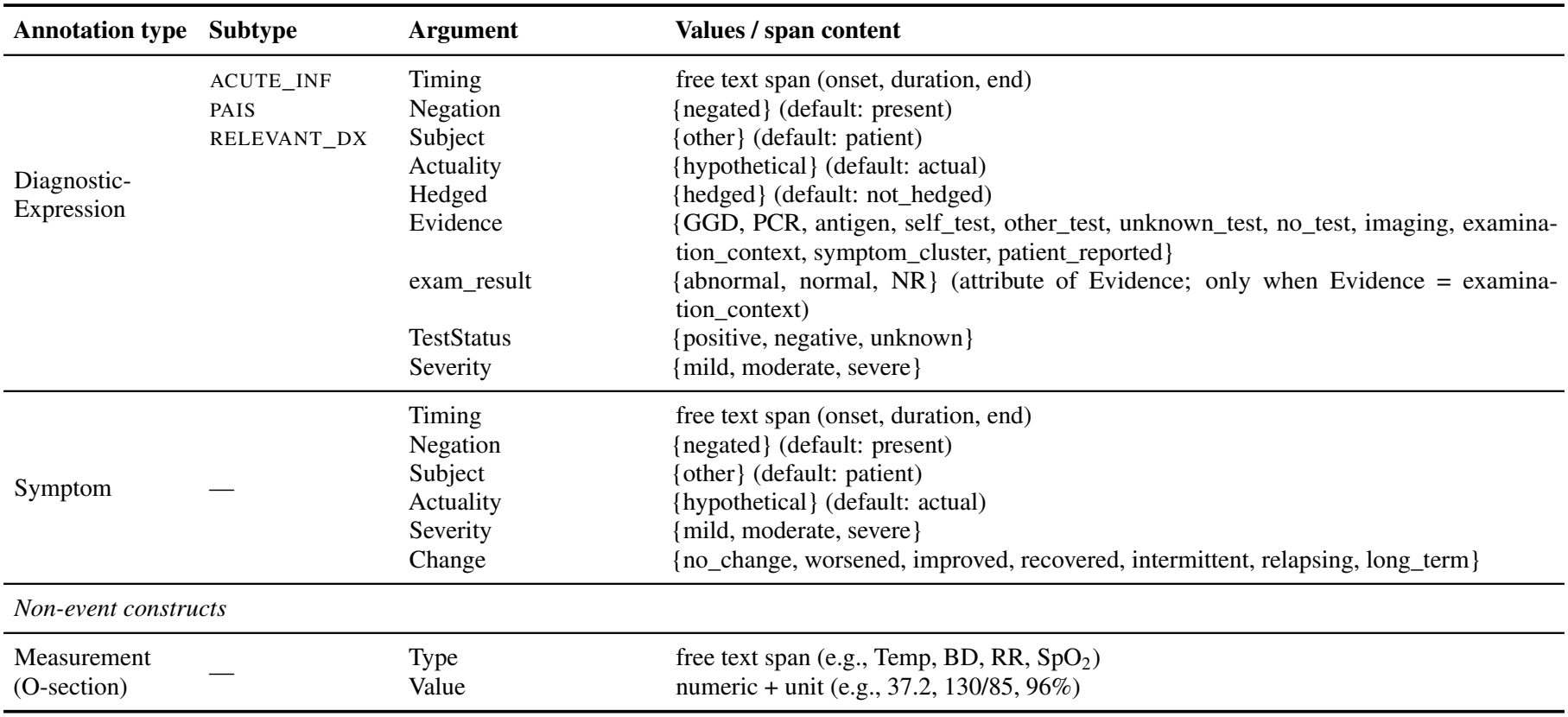
The annotation schema. Triggers anchor the annotation; arguments capture context and modality. ACUTE_INF: acute infection (COVID-19, influenza, RSV, other viral respiratory). PAIS: post-acute infection syndrome (post-COVID, ME/CVS, POTS, post-influenza). RELEVANT_DX: comorbidity or differential diagnosis. Measurements are standalone and cannot be negated.

| Annotation type | Subtype | Argument | Values / span content |
|---|---|---|---|
| Diagnostic-Expression | ACUTE_INF<br>PAIS<br>RELEVANT_DX | Timing | free text span (onset, duration, end) |
|  |  | Negation | {negated} (default: present) |
|  |  | Subject | {other} (default: patient) |
|  |  | Actuality | {hypothetical} (default: actual) |
|  |  | Hedged | {hedged} (default: not_hedged) |
|  |  | Evidence | {GGD, PCR, antigen, self_test, other_test, unknown_test, no_test, imaging, examina-tion_context, symptom_cluster, patient_reported} |
|  |  | exam_result | {abnormal, normal, NR} (attribute of Evidence; only when Evidence = examina-tion_context) |
|  |  | TestStatus | {positive, negative, unknown} |
|  |  | Severity | {mild, moderate, severe} |
| Symptom | — | Timing | free text span (onset, duration, end) |
|  |  | Negation | {negated} (default: present) |
|  |  | Subject | {other} (default: patient) |
|  |  | Actuality | {hypothetical} (default: actual) |
|  |  | Severity | {mild, moderate, severe} |
|  |  | Change | {no_change, worsened, improved, recovered, intermittent, relapsing, long_term} |
| *Non-event constructs* |  |  |  |
| Measurement<br>(O-section) | — | Type | free text span (e.g., Temp, BD, RR, SpO<sub>2</sub>) |
|  |  | Value | numeric + unit (e.g., 37.2, 130/85, 96%) |

**Figure 1.**
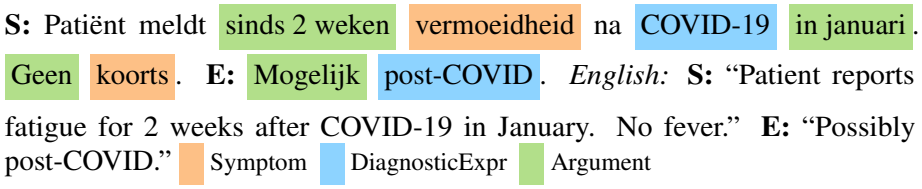
Annotated Dutch SOEP note (constructed example, no patient data). Triggers: fatigue (*vermoei-dheid*; Symptom, Timing=*sinds 2 weken*); COVID-19 (DiagExpr, ACUTE_INF, Timing=*in januari*); fever (*koorts*; Symptom, Negation=*Geen*); post-COVID (Di-agExpr, PAIS, Hedged=*Mogelijk*).

### 2.2 Adaptations from CACT

We retained the event-based architecture from CACT (Lybarger et al., 2021) and adapted the framework in four main areas. **(a) Diagnostic typology**: we introduced three DiagnosticEx-pression subtypes: acute infections, post-acute syndromes, and comorbidities/differential diagnoses. This supports the identification and comparison of post-infectious conditions across different infections and helps exclude alternative explanations. Diagnostic concepts were aligned with SNOMED CT (Donnelly, 2006), ICPC-1 (Lamberts and Wood, 1987) and UMLS (Bodenreider, 2004). **(b) Evidence granularity**: we introduced an Evidence argument with 11 values to capture the evidence supporting a diagnosis, distinguishing, for example, laboratory-confirmed infections from self-reported diagnoses. **(c) Temporal precision**: we extended the *Timing* annotation to capture the onset, duration and resolution of symptoms and diagnoses, allowing the extraction of temporal information relevant to PAIS classification, such as symptom onset or persistence at least 12 weeks after the acute infection. **(d) Context-dependent arguments**: we introduced *Subject* as a separate argument to distinguish conditions referring to the patient from those referring to others (e.g. ‘father has asthma’), which CACT captured within Assertion. We also distinguished *Hedged*, indicating uncer-tainty about a diagnosis being considered (e.g. ‘possibly post-COVID’, *mogelijk post-COVID*), from *Actuality*, indicating whether a condition is asserted as present or merely mentioned hypothetically (e.g. ‘afraid of long COVID’ or ‘to rule out bacterial superinfection’). This distinction is particularly relevant to GP documentation, where diagnostic uncer-tainty and the consideration of conditions not yet established play different roles in clinical reasoning and management. Safety-netting instructions in the Plan section are addressed by a separate annotation rule (see Appendix G).

### 2.3 Dutch Language Adaptations

#### Morphological complexity

Dutch compounding requires explicit trigger-boundary rules. For example, ‘common cold’ (*verkoudheid*) is annotated as a single trigger, while expressions such as *post-COVID-gerelateerde klachten* (‘post-COVID-related complaints’) require distinguishing the clinical concept from its contextual modifier. Anatomical modifiers specifying symptom location (e.g. *pijn in onderrug*, ‘lower back pain’) are included *in* the trigger span, while assertion modifiers (Hedged, Negation, Actuality) are annotated as separate arguments. **Clinical terminology variability**. Dutch GP notes contain a mixture of formal medical terminology, ICPC codes, and abbreviations (e.g. *BLWI* for upper respiratory tract infection, *gb* for *geen bijzonderheden*, ‘no abnormalities’). To ensure consistent annotation, the guide provides rules for interpreting common abbreviations and links each DiagnosticExpression subtype to established clinical terminologies. **Negation, hedging, and resolution**. Dutch clinical notes express negation and uncertainty with language-specific constructions, so annotation rules must account for their scope and meaning, and English heuristics such as NegEx (Chapman et al., 2001) cannot be reused directly. For example, *geen koorts* (‘no fever’) marks absence, whereas *geen koorts meer* (‘no fever anymore’) marks resolution. The guide therefore separates explicit absence (*geen koorts →* Negation{negated}), resolution (*geen koorts meer →*Change{recovered}), historical reference*→* ( Timing), and diagnostic uncertainty (*verdenking*,‘suspicion’ *→* Hedged{hedged}).

## 3 Annotation Methodology

### 3.1 Data and Annotation Process

We selected 200 SOEP notes from 200 patients within the Julius General Practitioners’ Network (JGPN; Smeets et al., 2018). Patients were selected based on ICPC-1 codes for acute respiratory infection, PAIS, or PAIS-related symptoms, without additional screening of the clinical notes (see sampling details in Appendix B). Two medical students independently annotated all 200 notes using INCEpTION (Klie et al., 2018) after structured training. Annotators first identified event triggers and subsequently annotated their contextual arguments. Calibration sessions were conducted to discuss annotation rules, identify ambiguities, and refine the annotation guideline. Following these sessions, both annotators revised their annotations using the updated guideline, without access to each other’s annotations. Disagreements were not resolved jointly at this stage. IAA was calculated on these final independent annotations, *before* adjudi-cation by three supervisors. The annotation process required approximately 15 hours per annotator. Both annotators were medical students employed as supervised research assistants and compensated according to standard institutional arrangements.

### 3.2 Agreement Metrics

We computed IAA using three metrics: **(i)** token-level Cohen’s *κ* for type-presence and type+subtype; **(ii)** span-level *F*_1_ using the matching rule applied to evaluate CACT, here termed the Lybarger criterion (Lybarger et al., 2021), requiring exact trigger spans, exact *(type, subtype)* agreement on labeled arguments, and token-level partial overlap on span-only arguments, with clinical macro-subtypes for EVIDENCE (Appendix D); and **(iii)** *F*_1_ conditional on annotation overlap and type agreement (Hripcsak and Rothschild, 2005), restricted to spans marked by both annotators, thereby separating schema-level disagreement from differences in annotation volume. For example, suppose *A*_1_ annotates *vermoeidheid* (Symptom) with Timing *sinds 2 weken* and *COVID-19* (DiagnosticExpression), while *A*_2_ annotates *vermoeidheid* with Timing *2 weken* but does not annotate *COVID-19*. Under metric (i), the tokens of *vermoeidheid* count as agreement and those of *COVID-19* as disagreement. Under metric (ii), the Symptom trigger matches exactly, the Timing spans match through token overlap, and *COVID-19* is counted as a missed annotation. Under metric (iii), *COVID-19* is excluded because it was annotated by only one annotator, so the score reflects labelling agreement on shared annotations only. Confidence intervals (CIs) were obtained by document-level bootstrap with 1000 iterations. IAA was calculated on the full 200-note pilot. We grouped the eleven annotation layers into six *core* entities (Symptom, DiagnosticExpression, Negation, Evidence, Timing, Measurement), representing the minimum information needed to establish that a clinical event was mentioned, negated, evidenced, and timed, and five *context-dependent* phenomena (Actuality, Hedged, Severity, Subject, Change), which further qualify certainty, ownership, and trajectory once the core event is established. Agreement is reported separately for these two groups because, as Section 4.1 shows, they behave very differently, with guideline ambiguity concentrated in the context-dependent phenomena.

## 4 Results

### 4.1 Inter-Annotator Agreement

**(i) Token-level Cohen’s** *κ*. Mean type-presence *κ* across the six core entities was 0.60 (Figure 2b, dark blue). Four of the eleven annotation layers showed substantial agreement (Landis and Koch, 1977) (Measurement *κ* = 0.80, Negation *κ* = 0.65, Timing *κ* = 0.63, and Subject *κ* = 0.64), four showed moderate agreement (DiagnosticExpression *κ* = 0.55, Change *κ* = 0.52, Evidence *κ* = 0.49, Symptom *κ* = 0.48), and three fell below 0.40 (Severity *κ* = 0.38, Hedged *κ* = 0.18, Actuality *κ* = 0.08). **(ii) Span-level** *F*_1_. Under the Lybarger matching criterion, aggregated span *F*_1_ across the six core entities was 0.51 [95% CI: 0.47, 0.55] when EVIDENCE was evaluated using macro subtypes (Figure 2b, medium blue). **(iii) Conditional** *F*_1_. Restricting to spans both annotators marked, conditional *F*_1_ across the six core entities was **0.78** (micro) [0.75, 0.80] (Figure 2b, light blue; per-entity values in Appendix C). **Inter-pretation**. The gap between span-level *F*_1_ (0.51) and conditional *F*_1_ (0.78) suggests that disagreement was mainly driven by annotation *coverage* rather than *label assignment* for shared events: *A*_2_ produced ~36% more annotations than *A*_1_, concentrated in EVIDENCE (Figure 2a), reflecting a broader inclusion criterion for examination-context evidence. Taking *A*_1_’s annotations as the reference set, *A*_2_ recovered 0.82 [95% CI 0.79, 0.84] of *A*_1_’s core-entity annotations; conversely, *A*_1_ recovered 0.75 [95% CI 0.73, 0.79]. EVIDENCE remained the most challenging core entity after conditioning on shared annotations (conditional *F*_1_ = 0.68), largely because of subtype variability within EXAMINATION_CONTEXT. By contrast, the low conditional *F*_1_ values for Actuality (0.16) and Hedged (0.39) indicate disagreement on label assignment rather than on annotation coverage. These findings suggest genuine annotation difficulty, motivating the planned calibration rounds to refine both the guideline and annotator training.

**Figure 2.**
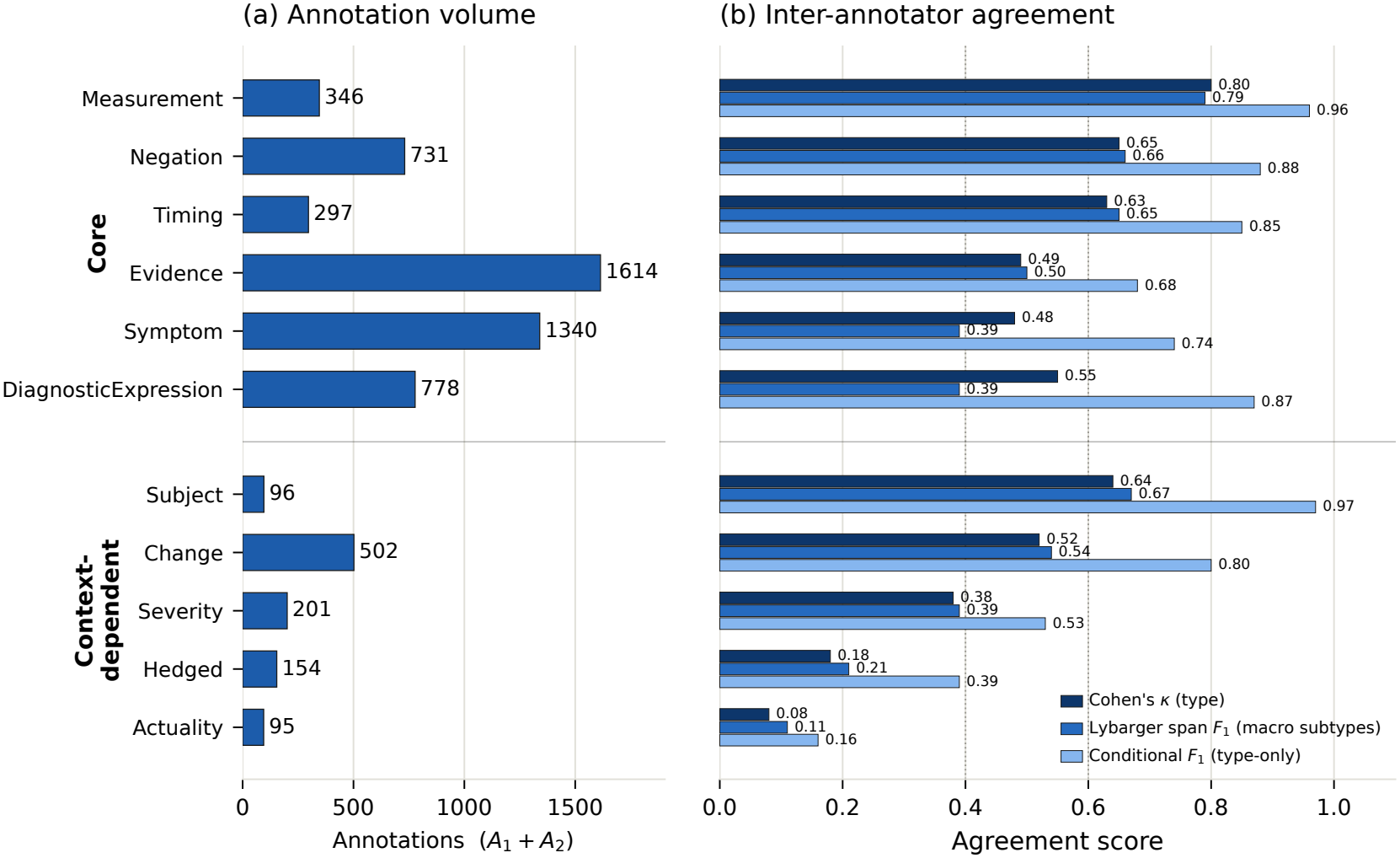
Pilot corpus distribution and inter-annotator agreement per layer. **(a)** Combined *pre-adjudication* annotation counts (raw union of both annotators’ spans, *A*_1_+*A*_2_, over the 200 notes; not a reconciled gold corpus). (a)Per-layer Cohen’s *κ* (dark blue), Lybarger span *F*_1_ with macro EVIDENCE subtypes (medium blue), and conditional *F*_1_ (Hripcsak and Rothschild, 2005) (light blue); core (top) and context-dependent (bottom) layers each ordered by Lybarger *F*_1_. Matching modes: Appendix C.

### 4.2 Corpus Statistics

Figure 2a shows the combined pre-adjudication annotation counts across the 200-note pilot (*A*_1_+*A*_2_). **Triggers**. Symptom was the most frequent trigger type, with 1340 combined annotations (3.6 and 3.1 per note for *A*_1_ and *A*_2_), consistent with the symptom-oriented structure of SOEP notes. Of the 778 DiagnosticExpression triggers, ACUTE_INF and RELEVANT_DX together accounted for 94.7%, whereas only 3.5% were labelled PAIS. This scarcity of explicit PAIS diagnoses supports the need for an extractor capable of identifying post-infectious phenotypes from *symptom constellations* rather than relying on explicit diagnostic labels alone. **Arguments**. EVIDENCE was the most frequent argument, with 1614 combined annotations. Disagreement on its `exam_result` attribute was concentrated in missing values: the attribute was left unspecified in 16% of cases for *A*_1_ versus 48% for *A*_2_ (Appendix F).

## 5 Conclusion and Future Work

We present a Dutch adaptation of CACT for event extraction from primary care records, covering symptoms, acute infections, diagnoses and contextual information, with specific extensions to support PAIS research. The released annotation guideline documents both the adapted schema and the Dutch-specific rules developed through pilot annotation and adjudication. Among the core layers, pilot IAA was highest for Measurement, Negation, and Timing, whereas the context-dependent layers Actuality, Hedged, and Severity showed sub-stantially lower agreement and are therefore prioritised for further guideline refinement and annotator calibration. Some elements of the adaptation are likely to transfer to other languages and diseases: the event-based backbone, the split between uncertain assertion (Hedged) and non-assertion (Actuality), and the SOEP-aware rules for negation and resolution. Others are specific to Dutch or to PAIS: the trigger-boundary rules for Dutch compounds, the handling of Dutch GP abbreviations, and the operationalisation of PAIS-specific temporal criteria through Timing spans. Whether this distinction holds across settings remains to be tested through adaptation to other languages or disease domains. The next phase will scale annotation to approximately 2,000 notes from four Dutch GP networks. These data will be used to train a Dutch clinical event extractor by fine-tuning MedRoBERTa.nl (Verkijk and Vossen, 2025), with comparison against a general-domain Dutch language baseline (Delobelle et al., 2020) and a translate-and-extract pipeline using a span-based extractor (Wadden et al., 2019).

## Limitations

### Single-region pilot

The pilot data were drawn from one primary-care network (JGPN). The planned rollout across four Dutch primary-care networks will provide a broader test of the schema and its generalisability across documentation settings.

### Single adaptation case

This study evaluates one adaptation of CACT: to Dutch primary care and to the PAIS domain. The proposed distinction between broadly transferable and case-specific adaptations (Section 5) is therefore hypothesis-generating rather than established. Testing it will require adaptation to additional languages or diseases.

### Annotation-volume asymmetry

The two annotators differed by ~36% in annotation volume, with most of the difference concentrated in EVIDENCE and attributable to differing inclusion criteria for examination-context evidence. We therefore report both span *F*_1_ and the conditional *F*_1_ (Hripcsak and Rothschild, 2005) to disentangle schema-level disagreement from this volume effect. A calibration round is planned to address both this volume asymmetry and the context-dependent phenomena with *κ <* 0.40 before annotation is scaled further.

### Annotator pool

Agreement estimates are based on two supervised medical students rather than practising GPs, and *n* = 2 provides limited information on how consistently the schema can be applied across annotators. The planned scale-up will include greater annotator diversity and continued clinical supervision.

### Context-dependent phenomena

Severity, Hedged and Actuality showed low agreement (*κ <* 0.40), indicating that their current definitions and decision rules require further refinement. Initial extractor development will therefore prioritise the six core entities, while the context-dependent phenomena are revised and re-evaluated in a subsequent calibration round.

## Ethical Considerations

This study does not fall under the scope of the Dutch Medical Research Involving Human Subjects Act (WMO) and therefore does not require approval from an accredited medical ethics committee in the Netherlands. At UMC Utrecht, an independent quality check was carried out to ensure compliance with legislation and regulations (informed consent procedure, data management, privacy and legal aspects), and the study is listed in the UMC Utrecht research register under number 25U-0459. Annotation runs in a secure INCEpTION instance within a protected environment; the released guideline uses only constructed examples and complies with GDPR. We release, with a persistent identifier, our guideline, an INCEpTION project export with the schema, and 500 synthetic SOEP notes with machine-generated annotations covering every value in Table 1. The synthetic notes are produced by a rule-based generator and are neither derived from nor paraphrased from real records. The generator samples four note templates (full SOEP, free paragraph, follow-up, and S and P only) and inserts Dutch GP abbreviations, ICPC-1 codes, date formats, typos, negation and hedging; the annotations are generated by rules from the same templates. The synthetic notes are therefore not a human-annotated gold set: the human annotation decisions are documented in the guideline and the schema export. Notes authored by clinicians about fictitious patients (Le Guellec et al., 2026; Tannier et al., 2026) would be a more realistic shareable alternative, which we leave to future work. The released artifacts (annotation guideline, INCEp-TION schema export, and synthetic notes) are our own work and are distributed under a permissive open licence (CC BY 4.0); the CACT framework is adapted conceptually and no CACT data is redistributed. The underlying GP corpus is not openly releasable under GDPR; however, data from the participating primary-care network (JGPN) may be requested for research purposes, subject to approval by the network’s research committee and compliance with the applicable data-use conditions. The released artifacts are available at https://doi.org/10.5281/zenodo.24033418.

## Data Availability

The underlying GP corpus is not openly releasable under GDPR; however, data from the participating primary-care network may be requested for research purposes, subject to approval by the network's research committee and compliance with the applicable data-use conditions. The annotation guideline, the annotation schema and the set of rule-generated synthetic notes produced in this work are openly available under CC BY 4.0.

## A Full Annotation Schema

Table 1 gives the complete annotation schema described in prose in the Schema Overview (Section 2.1).

## B Sampling Procedure

For each of the 200 pilot patients one SOEP note was extracted within *±*3 months of an index date: for 100 patients a note sampled uniformly at random, for the other 100 the longest note with all four SOEP sections. The 200 pilot patients were partitioned into two pools of 100 by alternating index assignment over the sorted patient list (seed = 42). For each patient, candidate SOEP notes were restricted to a *±*3-month window around the index date `fdate`; notes prior to 2008 were discarded. In the random pool, one note was drawn uniformly from the eligible window. In the longest-complete pool, the longest note containing all four SOEP sections (S, O, E, P) was selected; if no complete note existed, the longest note in the window was used as a fallback. Agreement is consistent across the two pools of 100, so the deliberately skewed long-note sampling does not inflate the reported figures: computing Table 2 separately per stratum gives Core (micro) 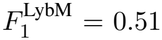(longest-complete) versus 0.48 (random) and 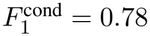 versus 0.81, both within the whole-pilot bootstrap CIs; reverse coverage (Cov *A*_2_ *→ A*_1_) is likewise stable (0.74 vs. 0.78).

**Table 2:** Inter-annotator agreement per entity under all matching criteria, computed on the raw, pre-adjudication annotations from both annotators over all 200 pilot notes. Conditional *F*_1_ and Cov are reported under the 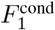 criterion. Bold rows are paper headline numbers. Cohen’s *κ* is at token-position level; all *F*_1_ values are span-level. Per-entity 95% bootstrap CIs are omitted for readability; micro-summary CIs are shown.

| Entity | κ (type) | κ (sub) | *F*<sub>1</sub><sup>strict</sup> | *F*<sub>1</sub><sup>Lyb</sup> | *F*<sub>1</sub><sup>LybM</sup> | *F*<sub>1</sub><sup>cond</sup> | Cov *A*<sub>1</sub> → *A*<sub>2</sub> | Cov *A*<sub>2</sub> → *A*<sub>1</sub> |
|---|---|---|---|---|---|---|---|---|
| *Core entities* |  |  |  |  |  |  |  |  |
| Symptom | 0.48 | 0.48 | 0.39 | 0.39 | 0.39 | 0.74 | 0.66 | 0.84 |
| DiagnosticExpr | 0.55 | 0.50 | 0.39 | 0.39 | 0.39 | 0.87 | 0.87 | 0.87 |
| Negation | 0.65 | 0.65 | 0.65 | 0.66 | 0.66 | 0.88 | 0.91 | 0.86 |
| Evidence | 0.49 | 0.48 | 0.16 | 0.52 | 0.52 | 0.68 | 0.92 | 0.55 |
| Timing | 0.63 | 0.63 | 0.54 | 0.65 | 0.65 | 0.85 | 0.76 | 0.96 |
| Measurement | 0.80 | 0.79 | 0.77 | 0.79 | 0.79 | 0.96 | 0.94 | 0.99 |
| **Core (micro)** | **0.60** | **0.57** | **0.39** | **0.51** | **0.51** | **0.78** | **0.82** | **0.75** |
| 95% CI |  |  | [0.37,0.45] | [0.48,0.56] | [0.47,0.55] | [0.75,0.80] | [0.79,0.84] | [0.73,0.79] |
| *Context-dependent phenomena (targets for guideline calibration)* |  |  |  |  |  |  |  |  |
| Subject | 0.64 | 0.58 | 0.48 | 0.62 | 0.67 | 0.97 | 0.97 | 0.97 |
| Change | 0.52 | 0.48 | 0.34 | 0.51 | 0.54 | 0.80 | 0.92 | 0.70 |
| Severity | 0.38 | 0.34 | 0.30 | 0.33 | 0.39 | 0.53 | 0.87 | 0.38 |
| Hedged | 0.18 | 0.18 | 0.19 | 0.21 | 0.21 | 0.39 | 0.30 | 0.55 |
| Actuality | 0.08 | 0.08 | 0.02 | 0.11 | 0.11 | 0.16 | 0.10 | 0.45 |
| **Context-dependent (micro)** | **0.36** | **0.33** | **0.30** | **0.40** | **0.43** | **0.66** | **0.70** | **0.62** |
| 95% CI |  |  | [0.21,0.31] | [0.32,0.42] | [0.34,0.45] | [0.53,0.66] | [0.69,0.81] | [0.55,0.68] |

## C Detailed Inter-Annotator Agreement Results

Table 2 reports IAA per entity type under all matching criteria evaluated:

- 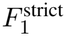exact span boundaries and exact sub-type match.
- 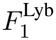Lybarger 2021 canonical (Lybarger et al., 2021): exact span for triggers, token overlap for arguments, exact subtype match.
- 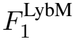Lybarger with macro subtypes (Appendix D). Single-valued subtype fields (e.g. Negation:negated) are auto-detected and treated as non-informative for matching.
- 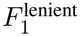token overlap, same entity type, subtype-free.
- 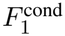conditional on annotation overlap (Hripcsak and Rothschild, 2005); see Appendix E.
- Cov *A*_1_ *→ A*_2_: fraction of *A*_1_’s spans that *A*_2_ replicated under 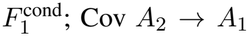 the reverse direction.

All 95% confidence intervals are obtained by document-level bootstrap with 1000 iterations.

## D Macro-Category Subtype Collapse

The 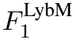 criterion collapses fine subtype values into clinical macro-categories before matching. The mappings follow our Annotation Guide and are clinically motivated rather than data-driven:

- **Evidence:source**: PCR, ANTIGEN, SELF_TEST, GGD, OTHER_TEST *→ lab_test*; IMAGING *imaging*; EXAMINATION_CONTEXT *→ clinical_exam*; PATIENT_REPORTED, SYMPTOM_CLUSTER *→ reported*; NO_TEST *→ no_test*; UNKNOWN_TEST *→ unknown*.
- **DiagnosticExpression:subtype** (INCEp-TION export labels; Table 1 abbreviates them): ACUTE_INFECTION *→ ACUTE*; PAIS, POST_COVID_PAIS*→ PAIS*; RELEVANT_DIAGNOSIS, *→* OTHER_DIAGNOSIS *COMORBID*.
- **Change:subtype**: NO_CHANGE, STABLE *→ STABLE*; IMPROVED, RECOVERED *→ IMPROVING*; WORSENED *→ WORSENING*; INTERMITTENT, RELAPSING *→ FLUCTUATING*; LONG_TERM *→ CHRONIC*.
- **Severity:subtype**: MILD *→ MILD*; MODERATE, SEVERE *→ MOD_HIGH*.

Single-valued subtype fields (Negation:negated, Hedged:hedged, Actuality:hypothetical, Subject:other) carry no information beyond the entity type and are auto-detected and excluded from the matching criterion under 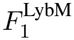and 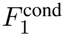. Despite EVIDENCE:SOURCE having eleven fine-grained values, 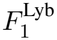and 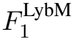 coincide for this layer (Table 2): annotator disagreement on SOURCE almost always crosses macro-category boundaries (e.g. IMAGING vs. EXAMINATION_CONTEXT) rather than occurring between fine values that share the same macro-category (e.g. PCR vs. ANTIGEN, both *lab_test*). The macro collapse is therefore clinically motivated but empirically inert for Evidence on this pilot; it is retained for consistency with DiagnosticExpression, Change, and Severity, where it does narrow the fine/macro gap (Table 2).

## E Conditional Agreement: Definition

Let *A*_1_ and *A*_2_ denote the span sets produced by the two annotators. Define

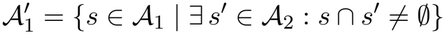

and analogously 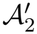 That is, 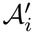contains only those spans that have at least one token overlap with *some* span in the other annotator’s set, regard-less of type, subtype, or exact boundary. 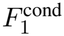 is then the standard lenient-type *F*_1_ computed on the pair 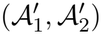 Intuitively, 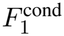 answers the question: *“conditional on both annotators having noticed something at this span position, did they agree on its type?”* Spans noticed by only one annotator are excluded from the denominator; such cases are not lost but reported separately as part of the asymmetric coverage view (Section 4.1 and Table 2). The formulation follows Hripcsak and Roth-schild (2005), who note that the *F*-measure is symmetric in its annotator inputs and that conditional-agreement framings are appropriate when annotators apply differing inclusion criteria, as is the case in our pilot.

## F Subtype Distributions

Table 3 reports the subtype distribution per entity, combining both annotators. Several patterns are informative for the downstream extractor design: PAIS accounts for only 3.5% of Diagnostic-Expression instances, confirming that PAIS phenotyping in primary care must rely on symptom constellations rather than explicit labels; Severity skews toward MILD (64%) consistent with the primary-care setting; Negation is almost exclusively NEGATED (99.9%), suggesting the subtype field is confirmatory and can be auto-detected (Appendix D); and the Evidence `exam_result` attribute shows substantial divergence in the MISSING-rate between annotators (16% for *A*_1_ vs. 48% for *A*_2_), the principal driver of the volume asymmetry reported in Section 4.1.

**Table 3:**
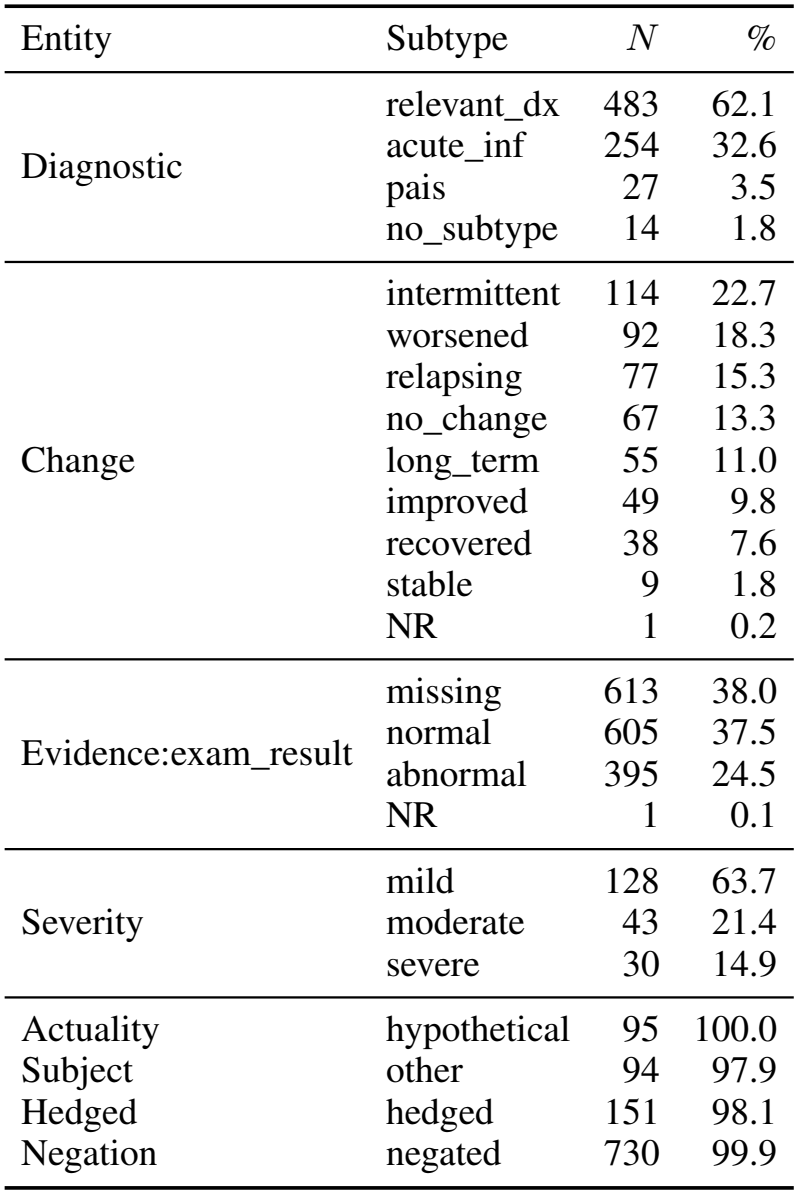
Combined-annotator subtype distribution (*A*_1_ + *A*_2_) on the 200-note pilot. DiagnosticExpression sub-types use the Table 1 names; the pilot export labels `other_diagnosis and post_covid_pais` cor-respond to RELEVANT_DX and PAIS. Entities with a single observed subtype value (Hedged, Negation, Actuality, Subject) are confirmatory-only fields and auto-detected by the IAA matching logic (Appendix D). Timing carries no subtype in the released schema. Measurement subtypes (`value/type`) are paired and not reported here. Evidence sub-arguments other than `exam_result` (`source, test_status`) are omitted for brevity; full breakdown available in the supplementary data.

| Entity | Subtype | *N* | *%* |
|---|---|---|---|
| Diagnostic | relevant_dx | 483 | 62.1 |
|  | acute_inf | 254 | 32.6 |
|  | pais | 27 | 3.5 |
|  | no_subtype | 14 | 1.8 |
| Change | intermittent | 114 | 22.7 |
|  | worsened | 92 | 18.3 |
|  | relapsing | 77 | 15.3 |
|  | no_change | 67 | 13.3 |
|  | long_term | 55 | 11.0 |
|  | improved | 49 | 9.8 |
|  | recovered | 38 | 7.6 |
|  | stable | 9 | 1.8 |
|  | NR | 1 | 0.2 |
| Evidence:exam_result | missing | 613 | 38.0 |
|  | normal | 605 | 37.5 |
|  | abnormal | 395 | 24.5 |
|  | NR | 1 | 0.1 |
| Severity | mild | 128 | 63.7 |
|  | moderate | 43 | 21.4 |
|  | severe | 30 | 14.9 |
| Actuality | hypothetical | 95 | 100.0 |
| Subject | other | 94 | 97.9 |
| Hedged | hedged | 151 | 98.1 |
| Negation | negated | 730 | 99.9 |

## G Guideline Excerpt: Hedged vs. Actuality

This excerpt from the released annotation guideline gives the decision rule for the two lowest-agreement context-dependent layers (Section 4.1).

All examples are constructed and contain no patient data. **Hedged**. Marks uncertainty or caution in a diagnosis. Default: {not_hedged} (certain). Example: *Mogelijk post-COVID; longembolie minder waarschijnlijk gezien lage risicofactoren*.

- *post-COVID* (DiagnosticExpression, PAIS), Hedged{hedged}: “Mogelijk”
- *longembolie* (DiagnosticExpression, RELEVANT_DIAGNOSIS), Hedged{hedged}: “minder waarschijnlijk”; Negation{negated}

### Hedged vs. Actuality

Hedged{hedged}: possible clinical explanation but uncertain whether it is true (“possibly influenza”, “suspected post-COVID”). Actuality{hypothetical}: mentioned without being asserted as a current clinical finding (“afraid of long COVID”, “to rule out bacterial superinfection”). Safety-netting instructions in the Plan section (e.g. “call back if worse, more short of breath, or coughing more”, *bij zieker worden / benauwd-heid / toename hoesten dan terugbellen*) are patient instructions, not current findings, and are not annotated. A hypothetical diagnosis to monitor for (“if worsening, consider pneumonia”, *bij verergering denken aan pneumonie*) is still annotated, with Actuality{hypothetical}.

